# Rebound in asthma exacerbations following relaxation of COVID-19 restrictions: a longitudinal population-based study (COVIDENCE UK)

**DOI:** 10.1101/2022.09.01.22279473

**Authors:** Florence Tydeman, Paul E Pfeffer, Giulia Vivaldi, Hayley Holt, Mohammad Talaei, David A Jolliffe, Gwyneth A Davies, Ronan A Lyons, Christopher J Griffiths, Frank Kee, Aziz Sheikh, Seif O Shaheen, Adrian R Martineau

**Affiliations:** Blizard Institute, Barts and The London School of Medicine and Dentistry, Queen Mary University of London, London, UK; William Harvey Research Institute, Barts and The London School of Medicine and Dentistry, Queen Mary University of London, London, UK; Department of Respiratory Medicine, Barts Health NHS Trust, London, UK; Wolfson Institute of Population Health, Barts and The London School of Medicine and Dentistry, Queen Mary University of London, London, UK; Population Data Science, Swansea University Medical School, Singleton Park, Swansea, UK; Asthma UK Centre for Applied Research, Queen Mary University of London, London, UK and University of Edinburgh, Edinburgh, UK; Centre for Public Health Research (NI), Queen’s University Belfast, Belfast, UK; Usher Institute, University of Edinburgh, Edinburgh, UK

**Author notes:** To whom correspondence should be addressed at the Blizard Institute, Barts and The London School of Medicine and Dentistry, Queen Mary University of London, 4 Newark St, London E1 2AT, UK. These authors contributed equally (Florence Tydeman, Paul E Pfeffer).

## Abstract

**Background:** The imposition of restrictions on social mixing early in the COVID-19 pandemic was followed by a reduction in asthma exacerbations in multiple settings internationally. Temporal trends in social mixing, incident acute respiratory infections (ARI) and asthma exacerbations following relaxation of COVID-19 restrictions have not yet been described.

**Methods:** We conducted a population-based longitudinal study in 2,312 UK adults with asthma between November 2020 and April 2022. Details of face covering use, social mixing, incident ARI and moderate/severe asthma exacerbations were collected via monthly on-line questionnaires. Temporal changes in these parameters were visualised using Poisson generalised additive models. Multilevel logistic regression was used to test for associations between incident ARI and risk of asthma exacerbations, adjusting for potential confounders.

**Results:** Relaxation of COVID-19 restrictions from April 2021 coincided with reduced face covering use (p<0.001), increased frequency of indoor visits to public places and other households (p<0.001) and rising incidence of COVID-19 (p<0.001), non-COVID-19 ARI (p<0.001) and moderate/severe asthma exacerbations (p=0.007). Incident non-COVID-19 ARI associated independently with increased risk of asthma exacerbation (adjusted odds ratio 5.75, 95% CI 4.75 to 6.97) as did incident COVID-19, both prior to emergence of the omicron variant of SARS-CoV-2 (5.89, 3.45 to 10.04) and subsequently (5.69, 3.89 to 8.31).

**Conclusions:** Relaxation of COVID-19 restrictions coincided with decreased face covering use, increased social mixing and a rebound in ARI and asthma exacerbations. Associations between incident ARI and risk of moderate/severe asthma exacerbation were similar for non-COVID-19 ARI and COVID-19, both before and after emergence of the SARS-CoV-2 omicron variant.

**Funding:** Barts Charity, UKRI

## INTRODUCTION

Asthma is an inflammatory airways disease that affects more than 300 million people globally.^1^ Episodes of progressive worsening of symptoms, termed exacerbations, are the major cause of morbidity and mortality in this condition. Viral respiratory infections are the most common precipitants of asthma exacerbations in both children and adults.^2^

Coronavirus disease 19 (COVID-19), an infectious disease caused by severe acute respiratory syndrome coronavirus 2 (SARS-CoV-2), emerged in Wuhan, China, in late 2019, and rapidly spread globally.^3^ In the absence of an effective vaccine or treatments, social distancing measures, such as lockdowns, were legislated globally. Imposition of these measures was associated with decreased social mixing, reduced transmission of SARS-CoV-2 and other acute respiratory pathogens ^4^ and reductions in asthma exacerbations.^5,6^ Although COVID-19 has been reported to associate with increased inhaler use and poorer symptom control in people with asthma,^17^ the relative influence of COVID-19 vs. non-COVID-19 ARI on risk of asthma exacerbation has yet to be evaluated. Moreover, there are limited data relating to whether relaxation of restrictions was associated with a rebound in acute respiratory infections (ARI) and asthma exacerbations.

A retrospective cohort study using data from an English national primary care database from January 2016 to October 2021 reported that asthma exacerbation rates were substantially lower between the second quarter of 2020 and the third quarter of 2021 than before the pandemic (2016-2019).^7^ Although this study did not formally demonstrate an increase in incidence of exacerbations over the course of 2021, a trend towards an increase in mean exacerbation rate from the first to the third quarter of 2021 was observed – a period which coincided with relaxation of COVID-19 restrictions in the UK. However, since the study was conducted using routinely collected data, it was not possible to establish whether changes in risk of asthma exacerbation over time were temporally associated with changes in behaviours affecting viral transmission. Moreover, follow-up did not extend to cover the period following emergence of the omicron variant of SARS-CoV-2, which became dominant in December 2021. This is an important knowledge gap, given concerns that the omicron variant may be more tropic to the upper airway and therefore potentially more likely than earlier variants to precipitate asthma exacerbations.^8^ We therefore sought to characterise changes in behaviours influencing respiratory viral transmission following relaxation of restrictions, and to establish whether changes in these behaviours coincided with increases in COVID-19, non-COVID-19 ARI and asthma exacerbations, using data from a prospective population-based longitudinal study of COVID-19 in the UK population (COVIDENCE UK).^9^ We also conducted multivariable analysis to determine the relative influence of COVID-19 and non-COVID-19 ARI on risk of asthma exacerbations, with additional stratification to determine whether the strength of association between COVID-19 and asthma exacerbations changed after the emergence of the omicron variant of SARS-CoV-2, as the dominant circulating variant in December 2021.

## METHODS

### Study Design and Setting and Approvals

COVIDENCE UK is a population-based longitudinal study investigating risk factors for, and impacts of, COVID-19 and other ARI. Full details of study design have been reported previously.^10-13^ Briefly, from 1st May 2020 to 6th October 2021, UK residents aged ≥16 years were invited via a national media campaign to complete a detailed online baseline questionnaire capturing self-reported information relating to their socio-demographic characteristics, occupation, lifestyle, quality of life, weight, height, longstanding medical conditions, medication use, vaccination status, diet and supplemental micronutrient intake. Follow-up on-line questionnaires capturing behaviours potentially influencing risk of acquiring or transmitting ARI, incident ARI symptoms, results of reverse transcription polymerase chain reaction (RT-PCR) and antigen testing for respiratory viruses, incident exacerbations of asthma and chronic obstructive pulmonary disease and doses of SARS-CoV-2 vaccine received were completed at monthly intervals. COVIDENCE UK is registered with ClinicalTrials.gov (ref NCT04330599) and was approved by Leicester South Research Ethics Committee (ref 20/EM/0117). All participants provided informed consent to participate, and 5-year follow-up with linkage to routinely collected medical record data is on-going.

### Participants

Inclusion criteria for the current analysis were participation in COVIDENCE UK; self-report of doctor-diagnosed asthma at baseline with on-going use of at least one prescribed asthma treatment and/or at least one moderate/severe asthma exacerbation (definition below) in the 12 months prior to enrolment; and completion of at least one monthly follow-up questionnaire between 12^th^ November 2020 (when details of asthma exacerbations were first included in monthly follow-up questionnaires) and 21^st^ April 2022 inclusive. Exclusion criteria were self-report of a positive RT-PCR or antigen test for SARS-CoV-2, on-going or prior chronic symptoms attributed to prior SARS-CoV-2 infection (long COVID) or hospitalisation for COVID-19 either prior to enrolment or at baseline.

### Variables

#### Behavioural Variables

Face covering use was categorised as the proportion of participants who reported that they had ‘Always (100% of the time)’ used a face covering while in an indoor public place in the week prior to questionnaire completion. Visits to shops, visits to other indoor public places, indoor visits to/from other households and public transport use were categorised as the proportion of participants who reported one or more such events in the week prior to questionnaire completion. Travel outside the UK was categorised as the proportion of participants who reported foreign travel in the month prior to questionnaire completion.

#### Respiratory Variables

Moderate/severe asthma exacerbations were defined as those requiring treatment with systemic corticosteroids and/or precipitating emergency department attendance or hospital admission. COVID-19 was defined as self-report of COVID-19 confirmed by a positive RT-PCR or antigen test result for SARS-CoV-2. Episodes of COVID-19 were imputed as being due to the omicron SARS-CoV-2 variant if they occurred from 16^th^ December 2021 onwards (i.e. the date on which the UK Health Security Agency reported omicron to be the dominant SARS-CoV-2 variant in the UK).^14^ Non-COVID-19 ARI was defined as self-report of a general practitioner (GP)or hospital diagnosis of pneumonia, influenza, bronchitis, tonsillitis, pharyngitis, ear infection, common cold or other upper or lower respiratory infection, and/or self-report of a symptom-defined ARI associated with at least one negative RT-PCR or antigen test results for SARS-CoV-2, and no positive RT-PCR or antigen test results for SARS-CoV-2. Symptom-defined episodes of ARI were identified using modified Jackson criteria for upper respiratory infection,^15^ modified Macfarlane criteria for lower respiratory infection^16^ and a triad of cough, fever and myalgia for influenza-like illness.^17^ Full details of algorithms employed are presented in Table S3, Supplementary Material.

#### Potential Confounders

The following factors were identified *a priori* as potential confounders of relationships between incident ARI and asthma exacerbation: duration of follow-up (number of months post enrolment), sex (male vs. female, defined by sex assigned at birth), age (years), ethnic origin (white vs. minority ethnic), current tobacco smoking (yes vs. no), educational attainment (primary/secondary school vs. higher/further education [A-levels] vs. college/university vs. post-graduate], body mass index (<25 vs. 25-30 vs. >30 kg/m^2^), asthma treatment level (use of reliever inhaler only vs. use of inhaled corticosteroid [ICS] +/-reliever inhaler vs. use of a long-acting bronchodilator inhaler +/-ICS or reliever inhaler vs. monoclonal antibody therapy with or without any other treatment), history of moderate/severe asthma exacerbation in the 12 months prior to enrolment (yes vs. no), index of multiple deprivation (IMD) 2019 score (assigned according to participants’ postcodes and categorised into quartiles), self-reported general health (poor vs. fair vs. good vs. very good vs. excellent) and SARS-CoV-2 vaccination status (primary vaccination course completed vs. not completed).

### Data sources and measurement

All variables above were derived from responses to baseline and monthly online questionnaires, detailed in Tables S1 and S2 of Supplementary Material, respectively.

### Bias

Selection bias was minimised by use of a national media campaign to enrol participants from the general population. Detection bias was minimised by using a uniform approach (online questionnaires) to capture outcomes in exposed vs. unexposed individuals. Observation bias was minimised as a result of participants being unaware of the specific hypotheses being tested in this analysis. Recall bias was minimised by frequent (monthly) administration of follow-up questionnaires.

### Study size

Details of the sample size calculation for the COVIDENCE UK study are presented elsewhere.^18^ The current analysis was pragmatic in nature, including all participants meeting the study-specific eligibility criteria described above with no sample size specified.

### Statistical Analysis

We used univariable Poisson generalised additive models to explore temporal changes in face covering use, social mixing behaviours, ARI and moderate/severe asthma exacerbations over time (November 2020 to April 2022). Univariable models were plotted to visualise the penalised regression splines over time (month-year), with transformation of *y*-axes onto the response scale. Means with 95% confidence intervals (CIs) of proportions experiencing each outcome were displayed. A fully adjusted multilevel logistic generalised linear model was applied to calculate adjusted odds ratios (aOR) for associations between incident ARI and moderate/severe asthma exacerbations. Adjusted models included age, sex, ethnicity, tobacco smoking status, educational attainment, body mass index, IMD 2019 quartile, self-defined general health, SARS-CoV-2 vaccination status, asthma treatment level and asthma exacerbation history prior to enrolment as covariates. Random effects of unique participant identifier were included in all models to account for within-participant variability. Missing data were assumed to be missing completely at random and were handled with listwise deletion in the generalised linear mixed models to obtain unbiased estimates. All statistical analyses were conducted using R version 4.1.1, with mixed effects models conducted using R-package lmer4.

## RESULTS

A total of 19,981 UK residents aged 16 years or more completed the COVIDENCE UK baseline questionnaire between 1^st^ May 2020 and 6^th^ October 2021, of whom 2,666 reported having doctor-diagnosed asthma with on-going use of a prescribed asthma treatment. Of these, 2,312 (86.7%) reported no episodes of COVID-19 before or at enrolment and completed at least one monthly follow-up questionnaire (Fig. 1). Table 1 presents baseline characteristics of participants contributing data to statistical analyses: 75.3% were female, median age was 60 years (interquartile range (IQR) 49-67), 94.7% were of White ethnic origin and 13.8% reported at least one moderate/severe asthma exacerbation in the 12 months prior to enrolment. Follow-up was from November 2020 to April 2022, during which 411 (17.8%) participants reported at least one moderate/severe asthma exacerbation, 656 (28.4%) reported at least one positive RT-PCR or antigen swab test result for SARS-CoV-2; and 1,115 (48.2%) reported at least one episode of ARI symptoms associated with a negative RT-PCR or antigen swab test result for SARS-CoV-2 (Table S4, Supplementary Material).

**Table 1.** Participant characteristics at baseline

|  |  |  |
| --- | --- | --- |
| Sex | Male, N (%) | 570 (24.7) |
|  | Female, N (%) | 1742 (75.3) |
| Age, years | Median (IQR) [range] | 60 (49–67) [16–88] |
| Ethnicity | Black/African/Caribbean/Black British, N (%) | 11 (0.5) |
|  | Mixed/multiple ethnic groups, N (%) | 34 (1.5) |
|  | South Asian,<sup>1</sup> N (%) | 43 (1.9) |
|  | White, N (%) | 2191 (94.7) |
|  | Other, N (%) | 33 (1.4) |
| Index of multiple deprivation, quartile<sup>2</sup> | 4 (least deprived), N (%) | 586 (25.4) |
|  | 3, N (%) | 587 (25.4) |
|  | 2, N (%) | 563 (24.4) |
|  | 1 (most deprived), N (%) | 570 (24.7) |
| Current tobacco smoking | Yes, N (%) | 123 (5.3) |
|  | No, N (%) | 2189 (94.7) |
| Educational attainment<sup>2</sup> | Primary/Secondary, N (%) | 229 (9.9) |
|  | Higher/Further (A levels), N (%) | 332 (14.4) |
|  | College/University, N (%) | 999 (43.2) |
|  | Post-graduate, N (%) | 748 (32.4) |
| Self-reported general health | Poor, N (%) | 148 (6.4) |
|  | Fair, N (%) | 450 (19.5) |
|  | Good, N (%) | 730 (31.6) |
|  | Very good, N (%) | 749 (32.4) |
|  | Excellent, N (%) | 235 (10.2) |
| Moderate/severe asthma exacerbation pre-enrolment<sup>3</sup> | Yes, N (%) | 320 (13.8) |
|  | No, N (%) | 1992 (86.2) |
| Asthma treatment<sup>2</sup> | Reliever only, N (%)<sup>4</sup> | 694 (30.0) |
|  | ICS +/- reliever, N (%)<sup>5</sup> | 686 (29.7) |
|  | LAB +/- ICS or reliever, N (%)<sup>6</sup> | 916 (39.6) |
|  | Monoclonal antibody, N (%)<sup>7</sup> | 14 (0.6) |

Abbreviations: ICS, inhaled corticosteroid. LAB, long-acting bronchodilator inhaler. 1, self-defined as Indian, Pakistani, Bangladeshi or Sri Lankan. 2, missing data: index of multiple deprivation (N=6), educational attainment (N=4), asthma treatment (N=2). 3, defined as reporting at least one asthma exacerbation requiring treatment with systemic corticosteroids and/or precipitating emergency department attendance or hospital admission in the 12 months preceding enrolment to COVidence UK. 4, inhaled short-acting beta-agonist or anticholinergic. 5, regular use of ICS +/- reliever inhaler, but no LAB or monoclonal antibody treatment. 6, regular use of LAB +/- ICS or reliever inhaler, but no monoclonal antibody treatment. 7, monoclonal antibody treatment, with or without any other treatment.

**Figure 1:**
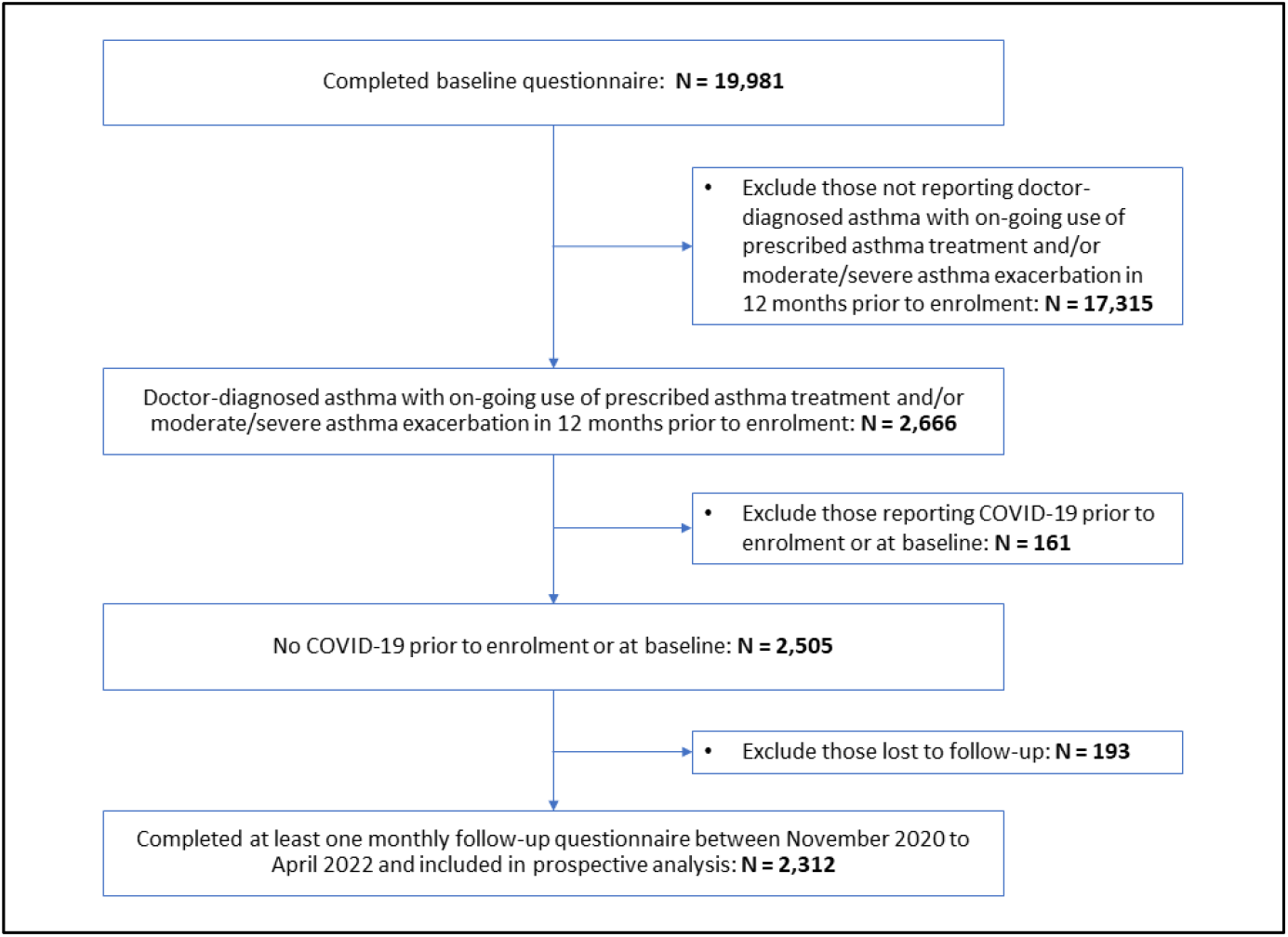
Participant Flow.

Figure 2 presents generalised additive model plots illustrating changes in a range of behaviours potentially influencing risk of acquiring or transmitting ARI over the study period. Statistically significant temporal fluctuations were seen in proportions of participants reporting face covering use (p<0.001), visits to shops (p<0.001), visits to other indoor public places (p<0.001), making or receiving indoor visits to or from other households (p<0.001), using public transport (p<0.001) or travelling outside of the UK (p<0.001). The period during which more stringent restrictions were imposed (November 2020 to March 2021) coincided with more frequent use of face coverings and reduced social mixing behaviours. Conversely, periods during which restrictions were relaxed (April to November 2021 and February to April 2022) coincided with less frequent face covering use and increased social mixing behaviours. The period immediately following emergence of omicron as the dominant variant of SARS-CoV-2 in the UK (December 2021 to January 2022) coincided with increased face covering use, reduced public transport use and less frequent travel outside the UK.

**Figure 2:**
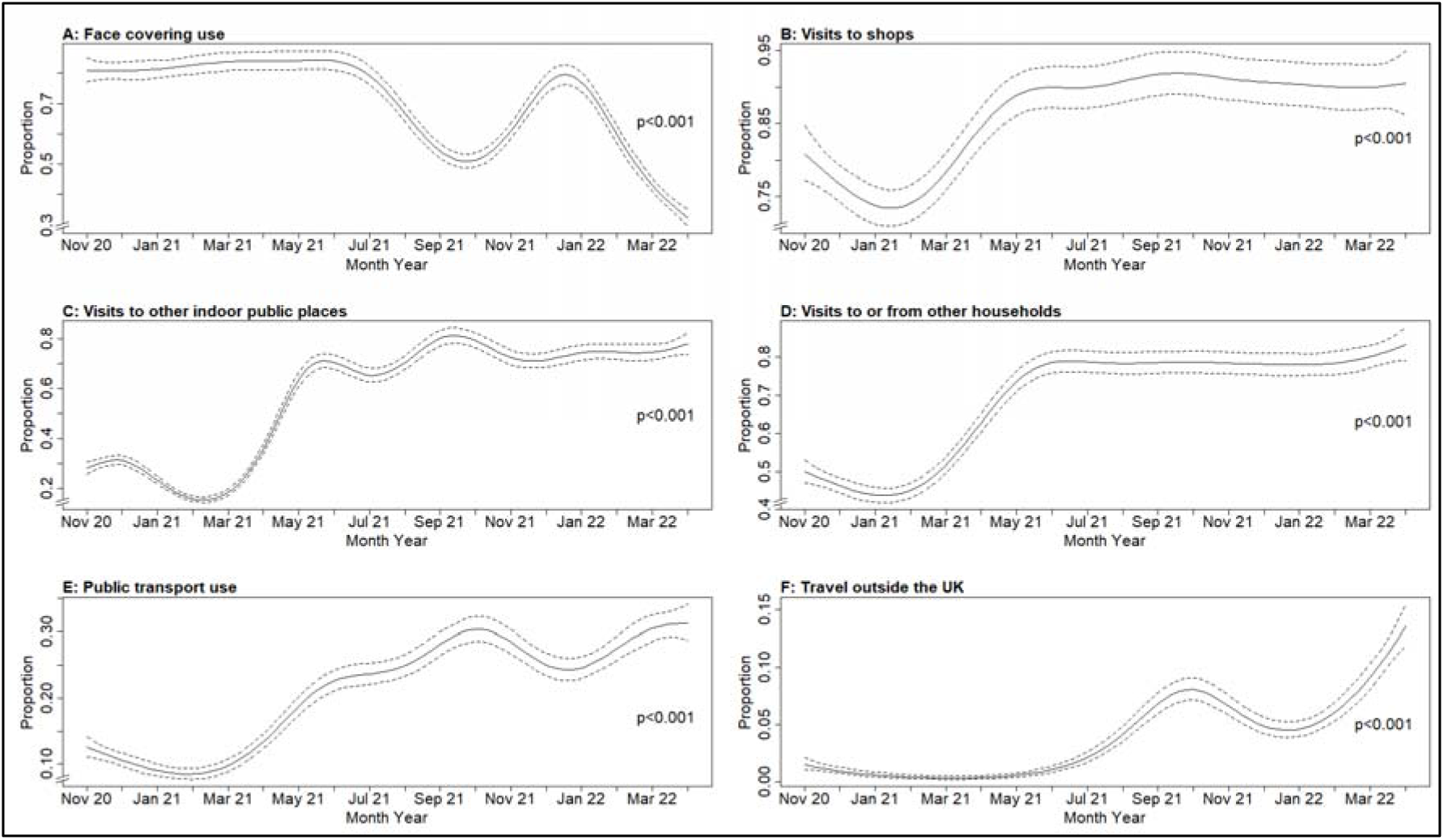
Temporal trends in behaviours potentially influencing risk of acquiring or transmitting acute respiratory infections from November 2020 to April 2022. **A**, proportion of participants using a face covering 100% of the time when visiting an indoor public place in the week prior to completion of monthly follow-up questionnaire. **B**, proportion of participants visiting shops at least once in the week prior to completion of monthly follow-up questionnaire. **C**, proportion of participants visiting an indoor public place other than a shop at least once in the week prior to completion of monthly follow-up questionnaire. **D**, proportion of participants making or receiving at least one indoor visit to another household in the week prior to completion of monthly follow-up questionnaire. **E**, proportion of participants using public transport at least once in the week prior to completion of monthly follow-up questionnaire. **F**, proportion of participants travelling outside the UK at least once in the month prior to completion of monthly follow-up questionnaire. All plots are from univariable generalised additive models; P values refer to temporal changes in proportions over the study period. Solid and dotted lines show means and 95% confidence intervals, respectively.

Figure 3 presents generalized additive model plots illustrating changes in incident ARI and asthma exacerbations over the study period. Statistically significant temporal fluctuations were seen in proportions of participants reporting COVID-19 (p<0.001), non-COVID-19 ARI (p<0.001) and asthma exacerbations (p=0.007). The proportion of participants reporting COVID-19 at each monthly questionnaire began to increase from June 2021, climbing more steeply from December 2021. By contrast, the proportion of participants reporting non-COVID-19 ARI increased from March to November 2021 but dipped from December 2021 onwards. The proportion of participants reporting asthma exacerbations increased more gradually from March 2021 and levelled off from January 2022 onwards.

**Figure 3:**
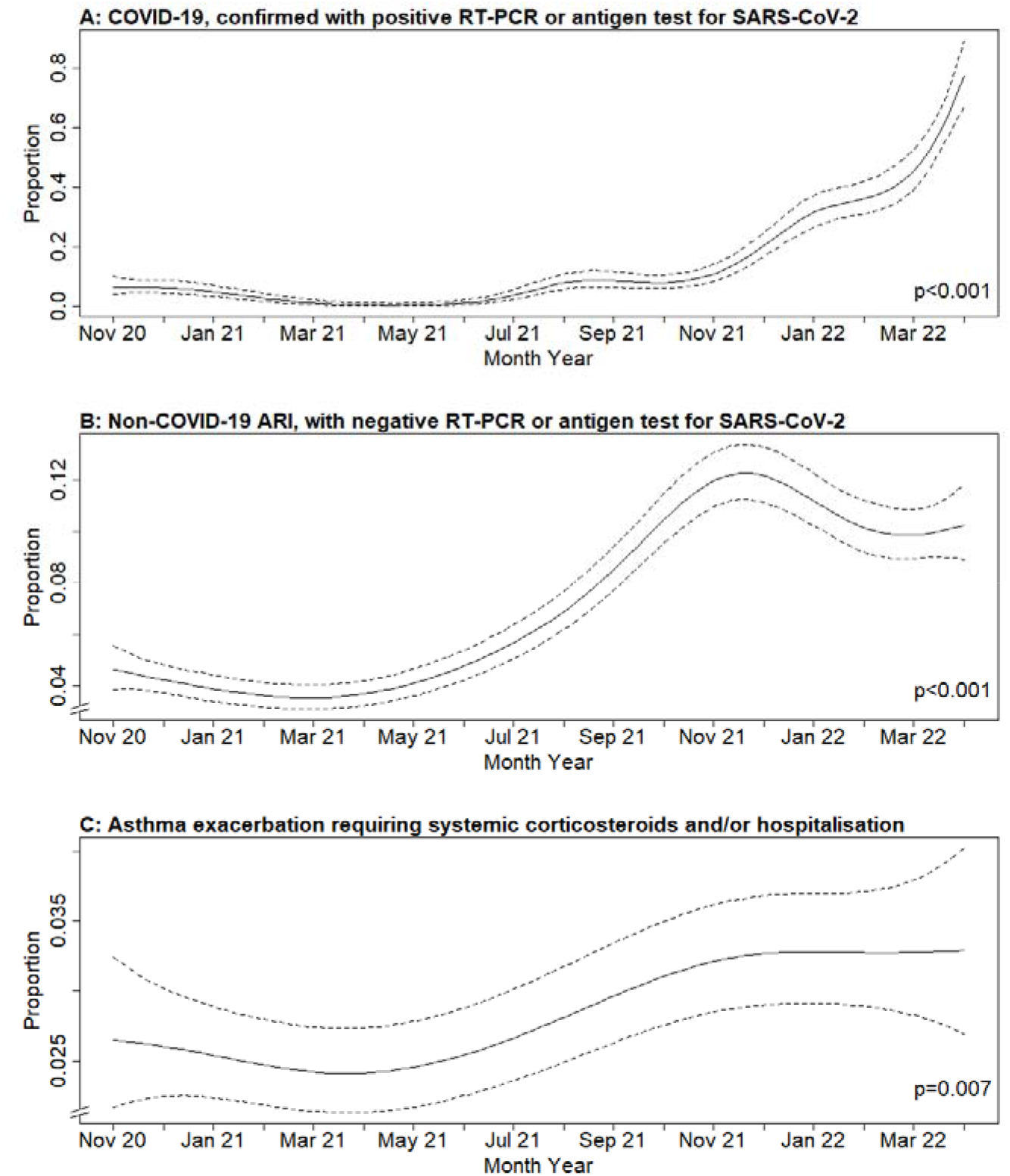
Temporal trends in incident acute respiratory infections and asthma exacerbations from November 2020 to April 2022. **A**, RT-PCR- or antigen test-confirmed COVID-19. **B**, episodes of acute respiratory infection symptoms associated with a negative RT-PCR or antigen test result for SARS-CoV-2. **C**, episodes of moderate/severe asthma exacerbation. All plots are from univariable generalised additive models; P values refer to temporal changes in proportions over the study period. Solid and dotted lines show means and 95% confidence intervals, respectively.

Table 2 presents results of univariable and multivariable analysis of potential determinants of moderate/severe asthma exacerbation. Incident non-COVID-19 ARI associated with increased odds of reporting an asthma exacerbation after adjustment for multiple potential sociodemographic and clinical confounders (aOR 5.75, 95% CI 4.75 to 6.97). Similarly, incident COVID-19 associated with increased odds of moderate/severe asthma exacerbation, both prior to dominance of the omicron variant of SARS-CoV-2 in December 2021 (aOR 5.89, 95% CI 3.45 to 10.04) and subsequently (aOR 5.69, 95% CI 3.89 to 8.31). Independent associations with increased odds of reporting an asthma exacerbation were also seen for participants receiving more intensive asthma treatment vs. use of a reliever inhaler only (for long-acting bronchodilator therapy, aOR 1.94, 95% CI 1.36 to 2.78; for monoclonal antibody asthma therapy, aOR 7.24, 95% CI 2.36 to 22.18); for those who reported an asthma attack in the 12 months prior to enrolment vs. those who did not (aOR 5.01, 95% CI 3.67 to 6.85); and for those who reported less good general health (‘good’ vs. ‘excellent’, aOR 1.99, 95% CI 1.04 to 3.80; ‘fair’ vs. ‘excellent’, aOR 3.17, 95% CI 1.64 to 6.16; ‘poor’ vs. ‘excellent’, aOR 4.57, 95% CI 2.19 to 9.55).

**Table 2:** Determinants of risk of moderate/severe asthma exacerbation

|  |  | Unadjusted OR<br>(95% CI) | P | Adjusted OR (95%<br>CI)<sup>1</sup> | P |
| --- | --- | --- | --- | --- | --- |
| Follow-up duration, months |  | 1.03 (1.02 to 1.05) | <0.001 | 0.99 (0.97 to 1.02) | 0.438 |
| Sex | Female | 1.00 (Ref) | - | 1.00 (Ref) | - |
|  | Male | 0.81 (0.58 to 1.12) | 0.205 | 1.11 (0.81 to 1.52) | 0.512 |
| Age, years |  | 0.99 (0.98 to 1.00) | 0.072 | 1.01 (1.00 to 1.02) | 0.202 |
| Ethnicity | White | 1.00 (Ref) | - | 1.00 (Ref) | - |
|  | Other | 1.01 (0.53 to 1.91) | 0.976 | 1.25 (0.69 to 1.52) | 0.461 |
| Current tobacco smoking | No | 1.00 (Ref) | - | 1.00 (Ref) | - |
|  | Yes | 1.96 (1.11 to 3.45) | 0.020 | 1.34 (0.78 to 2.31) | 0.290 |
| Educational attainment | Post-graduate | 1.00 (Ref) | - | 1.00 (Ref) | - |
|  | College/University | 1.32 (0.95 to 1.83) | 0.095 | 1.05 (0.77 to 1.42) | 0.774 |
|  | Higher/Further (A levels) | 1.57 (1.02 to 2.41) | 0.042 | 1.09 (0.72 to 1.64) | 0.688 |
|  | Primary/Secondary | 1.22 (0.74 to 2.02) | 0.442 | 1.06 (0.66 to 1.70) | 0.820 |
| Body mass index, kg/m<sup>2</sup> | <25 | 1.00 (Ref) | - | 1.00 (Ref) | - |
|  | 25-30 | 1.22 (0.87 to 1.70) | 0.253 | 1.12 (0.82 to 1.54) | 0.468 |
|  | ≥30 | 2.18 (1.57 to 3.03) | <0.001 | 1.36 (0.98 to 1.87) | 0.062 |
| Asthma treatment | Reliever only<sup>2</sup> | 1.00 (Ref) | - | 1.00 (Ref) | - |
|  | ICS +/- reliever<sup>3</sup> | 1.51 (1.02 to 2.24) | 0.041 | 1.24 (0.84 to 1.83) | 0.299 |
|  | LAB +/- ICS or reliever<sup>4</sup> | 3.67 (2.58 to 5.21) | <0.001 | 1.94 (1.36 to 2.78) | <0.001 |
|  | Monoclonal antibody<sup>5</sup> | 53.47 (16.39 to 174.46) | <0.001 | 7.24 (2.36 to 22.18) | 0.004 |
| Asthma exacerbation pre-enrolment<sup>6</sup> | No | 1.00 (Ref) | - | 1.00 (Ref) | - |
|  | Yes | 8.75 (6.55 to 11.70) | <0.001 | 5.01 (3.67 to 6.85) | <0.001 |
| Index of multiple deprivation, quartile | Q4 (Least Deprived) | 1.00 (Ref) | - | 1.00 (Ref) | - |
|  | Q3 | 0.96 (0.65 to 1.41) | 0.826 | 0.96 (0.66 to 1.38) | 0.807 |
|  | Q2 | 1.10 (0.75 to 1.63) | 0.618 | 1.04 (0.72 to 1.49) | 0.845 |
|  | Q1 (Most Deprived) | 1.35 (0.92 to 1.98) | 0.121 | 1.02 (0.71 to 1.47) | 0.900 |
| Self-reported general health | Excellent | 1.00 (Ref) | - | 1.00 (Ref) | - |
|  | Very good | 1.70 (0.91 to 3.19) | 0.096 | 1.73 (0.90 to 3.31) | 0.098 |
|  | Good | 2.85 (1.54 to 5.28) | 0.001 | 1.99 (1.04 to 3.80) | 0.037 |
|  | Fair | 5.90 (3.15 to 11.03) | <0.001 | 3.17 (1.64 to 6.16) | 0.001 |
|  | Poor | 12.23 (6.08 to 24.61) | <0.001 | 4.57 (2.19 to 9.55) | <0.001 |
| Primary course of SARS-CoV-2 vaccination | No | 1.00 (Ref) | - | 1.00 (Ref) | - |
|  | Yes | 1.35 (1.13 to 1.61) | 0.001 | 1.12 (0.84 to 1.48) | 0.466 |
| Incident COVID-19 | No | 1.00 (Ref) | - | 1.00 (Ref) | - |
|  | Yes, before Dec 2021 | 4.09 (2.22 to 7.53) | <0.001 | 5.89 (3.45 to 10.04) | <0.001 |
|  | Yes, on/after Dec 2021 | 4.17 (2.77 to 6.27) | <0.001 | 5.69 (3.89 to 8.31) | <0.001 |
| Incident non-COVID-19 ARI | No | 1.00 (Ref) | - | 1.00 (Ref) | - |
|  | Yes | 4.53 (3.72 to 5.52) | <0.001 | 5.75 (4.75 to 6.97) | <0.001 |

Abbreviations: ARI, acute respiratory infection; CI, confidence interval; ICS, inhaled corticosteroid; LAB, long-acting bronchodilator inhaler; OR, odds ratio. Ref, referent category. 1, mutually adjusted for all independent variables displayed. 2, inhaled short-acting beta-agonist or anticholinergic. 3, regular use of ICS +/- reliever inhaler, but no LAB or monoclonal antibody treatment. 4, regular use of LAB +/- ICS or reliever inhaler, but no monoclonal antibody treatment. 5, monoclonal antibody treatment, with or without any other treatment. 6, defined as reporting at least one asthma exacerbation requiring treatment with
systemic corticosteroids and/or precipitating emergency department attendance or hospital admission in the 12 months preceding enrolment to COVIDENCE UK.

## DISCUSSION

We report findings of the first study to compare the influence of COVID-19 vs. non-COVID-19 ARI on risk of asthma exacerbation, and to investigate the strength of association between COVID-19 and asthma exacerbation before and after emergence of the omicron variant of SARS-CoV-2. We show that relaxation of COVID-19 restrictions in the UK from April 2021 coincided with reduced use of face coverings, increased social mixing, and increases in COVID-19, non-COVID-19 ARI and moderate/severe asthma exacerbations. Incident ARI were strongly associated with increased risk of moderate/severe asthma exacerbations after adjustment for multiple potential confounders. However, the strength of such associations was similar for non-COVID-19 ARI and for COVID-19, both before and after emergence of the omicron variant of SARS-CoV-2.

Our findings support those of others who have investigated temporal trends in COVID-19 and asthma exacerbations over the course of the pandemic^5-7,19^ and extend them substantively by the addition of granular repeated-measures data capturing month-to-month changes in behaviours and non-COVID-19 ARI over an 18-month period spanning the lockdown of Winter 2020-21, the subsequent relaxation of restrictions and the emergence of the omicron variant of SARS-CoV-2. With regard to behaviours, our findings are broadly consistent with national data from the UK reporting good compliance with recommendations on use of coverings and reduced social mixing during early-2021, with increases in visiting and travel starting from April 2021.^20^ The temporal trends in COVID-19 reported here are also consistent with national data, which report increases in COVID-19 notifications from mid-2021, with sharp increases from November / December 2021,^21^ coinciding with the emergence of omicron as the dominant variant of SARS-CoV-2 in the UK.^22^ Similarly, national surveillance data show increases in cases of influenza, influenza-like illnesses and other respiratory viruses in 2021/2022 compared to 2020/2021, although incidence continued to remain lower than before the pandemic.^23^

Our findings also complement those of a retrospective cohort study conducted using data from an English national primary care database,^7^ which reported a trend towards an increase in mean asthma exacerbation rate over the course of 2021. Our demonstration that COVID-19 associates strongly with risk of moderate/severe exacerbation is in keeping with findings of a mixed-methods analysis reporting an association between COVID-19 and increased inhaler use in a cohort of adults with asthma.^24^ Given concerns that the omicron variant of SARS-CoV-2 may be more likely to precipitate asthma exacerbations than earlier variants,^8^ our finding that the strength of associations between COVID-19 and asthma exacerbation was not increased following emergence of the omicron variant is reassuring.

Our study has several strengths. The large size of our dataset, incorporating multiple repeated measures, afforded a high degree of power to detect temporal changes in behaviours and impacts of ARI on risk of asthma exacerbation. We captured episodes of non-COVID-19 ARI as well as COVID-19 across an extended period, which allowed us to compare the influence of different causes of ARI and different variants of SARS-CoV-2 on risk of asthma exacerbation. Availability of detailed information on multiple factors associating with exposures and outcomes of interest allowed us to perform adjusted analyses to minimise potential for confounding. The prospective longitudinal nature of the study allows us to rule out reverse causation as an explanation for associations observed, and the population-based approach to recruitment with inclusion of participants with different degrees of asthma severity enhances the generalisability of our findings.

Our study also has some limitations. No asthma outcome data were collected prior to November 2020, so information from before the pandemic and its early phases are lacking. All data are self-reported: however, participants were not aware of the hypotheses tested in these analyses, which reduces the risk of reporting bias. Variants of SARS-CoV-2 were imputed based on the date on which COVID-19 arose: this could lead to a degree of misclassification, although this is limited by the rapidity with which the omicron variant replaced the delta variant in the UK.^22^ We lacked information on participants’ adherence to asthma medication, which may have declined following relaxation of restrictions, and thereby contributed to subsequent increases in exacerbation risk. Our definition of non-COVID-19 ARI included symptom-defined events and was not validated against RT-PCR or other laboratory testing within this study; however, we have previously validated symptom-defined ARI against RT-PCR in a similar study population.^25^ Finally, we highlight that temporal associations between trends in behaviours, infections and asthma exacerbations should not necessarily be interpreted as being causal, given the observational nature of our study.

In conclusion, this large prospective population-based study shows for the first time that relaxation of COVID-19 restrictions in the UK coincided with increased risk of COVID-19 and non-COVID-19 ARI, which in turn associated independently with increased risk of moderate/severe asthma exacerbations. The strength of associations between incident ARI and asthma exacerbation were similar for non-COVID-19 ARI and COVID-19, both before and after emergence of omicron as the dominant variant of SARS-CoV-2 in the UK.

## Supporting information

STROBE checklist

## Data Availability

All data produced in the present study are available upon reasonable request to the authors, subject to terms of ethical approval.

## ACKNOWLEDGEMENTS

We thank all the people who participated in the COVIDENCE UK study, and the following organisations who supported study recruitment: Asthma UK, the British Heart Foundation, the British Lung Foundation, the British Obesity Society, Cancer Research UK, Diabetes UK, Future Publishing Ltd, Kidney Care UK, Kidney Wales, Mumsnet, the National Kidney Federation, the National Rheumatoid Arthritis Society, the North West London Health Research Register (DISCOVER), Primary Immunodeficiency UK, the Race Equality Foundation, SWM Health Ltd, the Terence Higgins Trust and Vasculitis UK.

## CONTRIBUTORS

FT, PEP and ARM conceived the study. HH co-ordinated and managed COVIDENCE UK, with input from ARM, MT, DAJ, GAD, RAL, CJG, FK, AS and SOS. FT, GV, HH, MT and DAJ contributed to data management. Statistical analyses were done by FT, with input from PEP and ARM. FT, PEP and ARM wrote the first draft of the report. All authors revised it critically for important intellectual content, gave final approval of the version to be published, and agreed to be accountable for all aspects of the work in ensuring that questions related to the accuracy or integrity of any part of the work were appropriately investigated and resolved.

## DISCLAIMER

The views expressed are those of the authors and not necessarily those of Barts Charity, BREATHE or Health Data Research UK.

## TRANSPARENCY DECLARATION

FT and ARM are the manuscript’s guarantors. They affirm that this is an honest, accurate, and transparent account of the study.

## DATA AVAILABILITY STATEMENT

Deidentified participant data are available from the corresponding author upon reasonable request, subject to the terms of Research Ethics Committee approval.

