## Supplementary Appendix for "Rebound in asthma exacerbations following relaxation of COVID-19 restrictions: a longitudinal population-based study (COVIDENCE UK)"

**Supplementary Material**

### **Table S1:** Baseline questionnaire

| **Sociodemographic** | |
| --- | --- |
| Date of questionnaire (DD/MM/YYYY) |  |
| Please state your **assigned sex at birth.** | -Male  -Female |
| Date of birth (DD/MM/YYYY) |  |
| What is your ethnic origin? | - White   - English / Welsh / Scottish / Northern Irish / British - Irish - Gypsy or Irish Traveller - Any other white background   - Mixed / Multiple ethnic groups   - White and Black Caribbean - White and Black African - White and Asian - Any other Mixed / Multiple ethnic backgrounds   - Asian / Asian British   - Indian - Pakistani - Bangladeshi - Chinese - Any other Asian background   - Black / African / Caribbean / Black British   - African - Caribbean - Any other Black / African / Caribbean background   - Arab  - Other Ethnic Group |
| Which of these best describes your use of cigarettes? | - I have never smoked cigarettes  - I used to smoke cigarettes occasionally but now not at all  - I used to smoke cigarettes daily but now not at all  - I smoke cigarettes occasionally but not every day  - I smoke cigarettes daily |
| What is the highest level of education that you have completed? | - Primary school  - Secondary school up to 16 years  - Higher or secondary or further education (A-levels, BTEC, etc.)  - College or university  - Post-graduate degree |
| What is your current weight? |  |
| What is your current height? |  |
| Have you ever been diagnosed with any of the following conditions by a doctor? | - Asthma  - Atopic Eczema or Atopic Dermatitis  - Autoimmune disease (e.g. rheumatoid arthritis, multiple sclerosis (MS), lupus (SLE), Crohn’s disease, ulcerative colitis, psoriasis, Raynaud’s disease, scleroderma)  - Cancer  - Cerebral Palsy  - COPD (including chronic bronchitis, and emphysema)  - Cystic Fibro  - Dementia  - Diabetes or pre-diabetes  - Hayfever or Allergic Rhinitis -  - Heart Attack, Angina or Coronary Artery Disease  - Heart Failure  - High Blood Pressure (Hypertension)  - HIV Infection  - Hyperparathyroidism (overactive parathyroid gland)  - Kidney stones  - Other kidney disease  - Leg Artery Disease (also known as ‘peripheral vascular disease’, ‘peripheral arterial disease’ or ‘intermittent claudication’)  - Mental health disorder  - Motor Neurone Disease  - Organ transplant  - Parkinson's Disease  - Primary immune deficiency (e.g. antibody deficiency, combined immunodeficiency)  - Sarcoidosis  - Sickle Cell Disease (i.e. two copies of altered gene, affected by anaemia and other complications  - Sickle Cell Carrier (also known as ‘sickle cell trait’, with only one copy of altered gene: few symptoms if any) -  - Splenectomy (removal of spleen)  - Stroke or Mini-Stroke  - Tuberculosis  - None of the above |
| If you have been diagnosed with Asthma, click as many as apply: | - I sometimes use a reliever inhaler (e.g. ventolin, salbutamol) to control my asthma symptoms  - I take a regular inhaler that includes a steroid preventer ONLY (e.g. Beclomethasone, Budesonide, Ciclesonide, Fluticasone or Mometasone)  - I take a regular inhaler that includes a long-acting bronchodilator ONLY (e.g. salmeterol, formoterol)  - I take a regular combination inhaler that contains BOTH a steroid preventer AND a long-acting bronchodilator (e.g. Seretide, Symbicort, Flutiform, Fostair)  - My asthma is being treated with monoclonal antibody infusions at the hospital  - In the last 12 months I have had one or more asthma attacks requiring treatment with steroid tablets (prednisolone)  - In the last 12 months I have had one or more asthma attacks requiring hospital admission |
| Postcode |  |
| Over the last 12 months, would you say that on the whole, your health has been: | - Excellent  - Very good  - Good  - Fair  - Poor |
| Have you had one or more doses of a COVID-19 vaccine (immunisation)? First doses and booster doses both count. | - Yes  - No  - Not sure |
| Since February 1st 2020, have you had a nose/throat swab to test for COVID-19? | - Yes  - No  - Not sure e.g. you took part in vaccine trial, but don’t yet know whether or not you had the real vaccine or the placebo (dummy vaccine) |

### **Table S2:** Monthly follow-up questionnaire

| **Questions asked at every monthly follow-up** |  |
| --- | --- |
| Since you last checked in with us, have you had an attack (b) of asthma or COPD (chronic bronchitis / emphysema)? | - Yes  - No |
| Was this an attack of asthma or COPD | - Asthma attack  - COPD attack |
| Did this asthma/COPD attack require treatment with steroid tablets (e.g. prednisolone)? | - Yes  - No  - Don’t know / not sure |
| What did the hospital doctors diagnose? Select as many as apply. | - Suspected or proven COVID-19  - Pneumonia  - ‘Flu' (influenza)  - Bronchitis  - Tonsillitis or pharyngitis (sore throat)  - Ear infection (otitis media)  - Common cold  - Another upper respiratory infection  - Another lower respiratory infection  - Asthma attack (flare-up or exacerbation of asthma symptoms)  - COPD attack (flare-up or exacerbation of COPD symptoms)  - Something else |
| Since you last checked in with us, have you had a nose or throat swab for COVID-19 or any other respiratory virus, or has a result from a previous swab test become newly available?(This question is about tests to detect the virus itself: they are usually done in somebody who has symptoms, but screening of asymptomatic people can also be done. It’s usually a nose/throat swab, but saliva tests are also becoming available) | - Yes  - No |
| On what date did you have this nose / throat swab? If you are not sure of the exact date, enter the approximate date (DD/MM/YYYY). |  |
| What was the result? Click as many as apply. | - Positive for COVID-19 (SARS-CoV-2 coronavirus)  - Positive for influenza virus  - Positive for another respiratory virus  - Negative for all/any viruses tested  - Not Known |
| What did the GP diagnose? Tick as many as apply | - Suspected or proven COVID-19  - Pneumonia  - ‘Flu' (influenza)  - Bronchitis  - Tonsillitis or pharyngitis (sore throat)  - Ear infection (otitis media)  - Common cold  - Another upper respiratory infection  - Another lower respiratory infection  - Something else |
| _1_How ill did you feel at your worst? | - Mildly unwell - I could do most of my usual activities  - Moderately unwell - I couldn’t do usual activities, but didn’t need to go to bed in the daytime (2)  - Very unwell – I had to go to bed in the daytime (3) |
| _2_Did you have a fever (high temperature)? | - Yes  - No |
| _3_Did you have a persistent cough (coughing a lot for more than an hour, or 3 or more coughing episodes in 24 hours)? | - No  - Persistent dry cough (i.e. producing little or no phlegm)  - Persistent productive cough |
| _4_Did you have a headache? | - Yes  - No |
| _5_Did you have muscle aches? | - Yes  - No |
| _6_Did you experience unusual shortness of breath? | - No  - Yes, mild symptoms - slight shortness of breath during ordinary activity  - Yes, significant symptoms – breathing was comfortable only at rest  - Yes, severe symptoms - breathing was difficult even at rest |
| _7_Please indicate any other symptoms you had | - Sore throat  - Sneezing  - Runny nose  - Blocked nose  - Unusually hoarse voice  - Unusual chest pain  - Unusual abdominal pain  - Diarrhoea  - Confusion, disorientation, or drowsiness  - Raised, red itchy welts on the skin or sudden swelling of the face or lips  - Red/purple sores or blisters on your feet or toes  - Unusual soreness or discomfort of the eyes (e.g. light sensitivity, excessive tears, pink/red eyes)  - Other symptom (please specify)  - None of the above |

### **Table S3:** Algorithms for symptom-defined episodes of non-COVID-19 acute respiratory infection (ARI).

Episodes fulfilling the criteria below were defined as non-COVID-19 ARI if they were associated with at least one negative RT-PCR or antigen test results for SARS-CoV-2, and no positive RT-PCR or antigen test results for SARS-CoV-2.

| **Non-COVID-19 ARI** | **Contributing questionnaire question (see Table S2 for subscript numbering):** | **Responses:** |
| --- | --- | --- |
| Upper respiratory infection | 1. How ill did you feel at your worst?  2. Did you have a fever (high temperature)?  3. Did you have a persistent cough (coughing a lot for more than an hour, or 3 or more coughing episodes in 24 hours)?  4. Did you have a headache?  7. Please indicate any other symptoms you had: | At least one nasal symptom   - 7. Sore throat OR Sneezing OR Runny nose   **AND**  At least one of the following symptoms:   - 1. Moderately unwell - I couldn’t do usual activities, but didn’t need to go to bed in the daytime OR Very unwell – I had to go to bed in the daytime   **OR**   - 2. Yes   **OR**   - 3. Persistent productive cough (i.e., producing little or no phlegm)   **OR**   - 4. Yes   **OR**   - 7. Sore throat OR unusually hoarse voice |
| Lower respiratory infection | 3. Did you have a persistent cough (coughing a lot for more than an hour, or 3 or more coughing episodes in 24 hours)?  6. Did you experience unusual shortness of breath?  7. Please indicate any other symptoms you had: | - 3. Persistent productive cough (i.e., producing little or no phlegm)   **OR**   - 3. Persistent dry cough + 7. Unusual chest pain   **OR**  3. Persistent dry cough + 6. Yes, mild symptoms - slight shortness of breath during ordinary activity OR yes, significant symptoms – breathing was comfortable only at rest OR Yes, severe symptoms - breathing was difficult even at rest |
| Influenza like illness (ILI) | 2. Did you have a fever (high temperature)?  3. Did you have a persistent cough (coughing a lot for more than an hour, or 3 or more coughing episodes in 24 hours)?  5. Did you have muscle aches? | - 2. Yes + 3. Persistent dry cough (i.e., producing little or no phlegm) OR Persistent productive cough + 5. Yes |

### **Table S4:** Proportion of participants reporting at least one episode of asthma exacerbation or acute respiratory infection during follow-up

| **Event** | **Proportion experiencing at least once** |
| --- | --- |
| Asthma exacerbation requiring treatment with systemic corticosteroids | 411/2312 (17.8%) |
| Incident COVID-19, confirmed with positive RT-PCR or antigen test for SARS-CoV-2 | 656/2312 (28.4%) |
| Incident non-COVID ARI, associated with negative RT-PCR or antigen test for SARS-CoV-2 | 1115/2312 (48.2%) |

ARI, acute respiratory infection. RT-PCR, reverse transcriptase polymerase chain reaction.
